# Clinical and molecular heterogeneity of pineal parenchymal tumors: a consensus study

**DOI:** 10.1101/2020.08.18.20172072

**Authors:** Anthony P.Y. Liu, Bryan K. Li, Elke Pfaff, Brian Gudenas, Alexandre Vasiljevic, Brent A. Orr, Christelle Dufour, Matija Snuderl, Matthias A. Karajannis, Marc K. Rosenblum, Eugene I. Hwang, Ho-Keung Ng, Jordan R. Hansford, Alexandru Szathmari, Cécile Faure-Conter, Thomas E. Merchant, Katja von Hoff, Martin Mynarek, Stefan Rutkowski, Felix Sahm, Cynthia Hawkins, Giles W. Robinson, Amar Gajjar, Stefan M. Pfister, Eric Bouffet, Paul A. Northcott, David T.W. Jones, Annie Huang

**Affiliations:** Department of Oncology, St. Jude Children’s Research Hospital, Memphis, TN, USA; Division of Hematology/Oncology, Department of Pediatrics, The Hospital for Sick Children, University of Toronto, Toronto, ON, Canada; Arthur and Sonia Labatt Brain Tumour Research Centre, Hospital for Sick Children, Toronto, ON, Canada; Laboratory Medicine and Pathobiology, Faculty of Medicine, University of Toronto, Toronto, ON, Canada; Hopp Children’s Cancer Center Heidelberg (KiTZ), Heidelberg, Germany; Pediatric Glioma Research Group (B360), German Cancer Research Center (DKFZ), Hopp Children’s Cancer Center Heidelberg (KiTZ), Heidelberg, Germany; Department of Pediatric Oncology, Hematology and Immunology, Heidelberg University Hospital, Heidelberg, Germany; Department of Developmental Neurobiology, St. Jude Children’s Research Hospital, Memphis, TN, USA; Faculté de Médecine, Université de Lyon, Lyon, France; Service d’Anatomie et Cytologie Pathologiques, CHU de Lyon, Lyon, France; Department of Pathology, St. Jude Children’s Research Hospital, Memphis, TN, USA; Département de Cancérologie de l’Enfant et de l’Adolescent, Institut Gustave Roussy, Villejuif, Paris, France; Division of Neuropathology, NYU Langone Health, New York, USA; Laura and Isaac Perlmutter Cancer Center, NYU Langone Health, New York, USA; Division of Molecular Pathology and Diagnostics, NYU Langone Health, New York, USA; Department of Pediatrics, Memorial Sloan Kettering Cancer Center, New York, USA; Department of Pathology, Memorial Sloan Kettering Cancer Center, New York, USA; Children’s National Medical Center, Washington, DC, USA; Department of Anatomical and Cellular Pathology, The Chinese University of Hong Kong, Shatin, New Territories, Hong Kong, China; Children’s Cancer Centre, The Royal Children’s Hospital; Murdoch Children’s Research Institute; Department of Pediatrics, University of Melbourne, Melbourne, VIC, Australia; Département de Neurochirurgie Adulte et Pédiatrique, Hôpital Femme Mère Enfant, Hospices Civils de Lyon, Bron, France; Institut d’Hématologie et d’Oncologie Pédiatrique, IHOPe, Lyon, France; Department of Radiation Oncology, St. Jude Children’s Research Hospital, Memphis, TN, USA; Department of Pediatric Oncology/Hematology, Charité-Universitätsmedizin Berlin, Berlin, Germany; Department of Paediatric Haematology and Oncology, University Medical Centre Hamburg-Eppendorf, Hamburg, Germany; Department of Neuropathology, Institute of Pathology, University Hospital Heidelberg, Heidelberg, Germany; Clinical Cooperation Unit Neuropathology, German Cancer Research Center (DKFZ), German Consortium for Translational Cancer Research (DKTK), Heidelberg, Germany; Division of Pathology, The Hospital for Sick Children, Toronto, ON, Canada; Division of Pediatric Neurooncology, German Cancer Research Center (DKFZ), Heidelberg, Germany; Department of Medical Biophysics, Faculty of Medicine, University of Toronto, Toronto, ON, Canada

**Keywords:** Pineoblastoma, pineal parenchymal tumors of intermediate differentiation, DNA methylation profiling, subgroup, consensus, miRNA, *KBTBD4*

## Abstract

**Background:** Recent genomic studies have shed light on the biology and inter-tumoral heterogeneity underlying pineal parenchymal tumors, in particular pineoblastomas (PBs) and pineal parenchymal tumors of intermediate differentiation (PPTIDs). Previous reports, however, had modest sample sizes and lacked power to integrate molecular and clinical findings. The different proposed subgroup structures also highlighted a need to reach consensus on a robust and relevant classification system.

**Methods:** We performed a meta-analysis on 221 patients with molecularly characterized PBs and PPTIDs. DNA methylation profiles were analyzed through complementary bioinformatic approaches and molecular subgrouping was harmonized. Demographic, clinical and genomic features of patients and samples from these pineal tumor subgroups were annotated.

**Findings:** Four clinically and biologically relevant consensus PB subgroups were defined: PB-miRNA1 (n=96), PB-miRNA2 (n=23), PB-MYC/FOXR2 (n=34) and PB-RB1 (n=25); with PPTID (n=43) remaining as a molecularly distinct entity. Genomic and transcriptomic profiling allowed the characterization of oncogenic drivers for individual subgroups, specifically, alterations in the microRNA processing pathway in PB-miRNA1/2, *MYC* amplification and *FOXR2* overexpression in PB-MYC/FOXR2, *RB1* alteration in PB-RB1, and *KBTBD4* insertion in PPTID. Age at diagnosis, sex predilection and metastatic status varied significantly among tumor subgroups. While patients with PB-miRNA2 and PPTID had superior outcome, survival was intermediate for patients with PB-miRNA1, and dismal for those with PB-MYC/FOXR2 and PB-RB1.

**Interpretation:** We systematically interrogated the clinical and molecular heterogeneity within pineal parenchymal tumors and proposed a consensus nomenclature for disease subgroups, laying the groundwork for future studies as well as routine use in tumor classification.

## Introduction

Pineal parenchymal tumors are rare central nervous system (CNS) neoplasms that encompass a spectrum of entities with varied histologic appearance and clinical phenotypes.^1^ Accounting for <1% of all CNS tumors, these range from World Health Organization (WHO) Grade 1 pineocytomas, to WHO Grade 2-3 pineal parenchymal tumors of intermediate differentiation (PPTIDs), and Grade 4 pineoblastomas (PBs), an embryonal tumor predominantly of pediatric onset.^2,3^ While resection alone will typically suffice for treatment of pineocytoma, the optimal adjuvant therapy needed for patients with PPTIDs is unclear, and cure is achieved in only two-third of patients with PB despite intensive cytotoxic therapy and radiotherapy in those eligible to be irradiated.^4^

Over the past few years, multi-omic profiling has enhanced our understanding of the biology that underlies pineal parenchymal tumors. In addition to loss of *RB1* function, alterations in microRNA (miRNA) processing genes, including loss-of-function mutations and deletions targeting *DICER1, DROSHA* and *DGCR8*, have emerged as prominent genetic events in PB.^5-9^ Indeed, PB represents the most common CNS malignancy for individuals with germline *DICER1* mutation, or DICER1 syndrome.^10^ Genome-wide DNA methylation studies have demonstrated the molecular distinctiveness of PB despite histological resemblance to other CNS embryonal tumors.^11^ Parallel independent studies have recently described inter-tumoral heterogeneity, with four to five molecularly distinct subgroups of PB.^5-7^ In addition to recurrent alterations in miRNA-processing genes and *RB1, FOXR2* overexpression and *MYC* amplification were observed in PBs from young children without other putative oncogenic drivers. Methylation analyses indicate PPTIDs are epigenetically different from PBs, with a majority exhibiting in-frame insertions in the Kelch domain of *KBTBD4*, a CUL3-based E3 ubiquitin ligase.^7,11,12^

Optimal treatment strategies for patients with PB and PPTID remain unknown, as clinical trials to date have accrued few of these rare tumors and grouped PB with other supratentorial CNS embryonal tumors.^13-16^ Current standard of care considers PB to be a high-risk pediatric embryonal brain tumor requiring maximal safe resection, followed by high dose craniospinal irradiation (CSI) with tumor boost and multi-agent chemotherapy, while strategies to delay or avoid radiotherapy have been used in very young children. Combined radio-chemotherapy approaches have also been extrapolated to the management of some PPTID, where clinical literature on optimal clinical care is scarce. Neurosurgical management of pineal parenchymal tumors is technically demanding.^17^ Gross tumor removal is often advocated on the basis of medulloblastoma studies, although evidence supporting the value of aggressive resection for patients with PB is inconsistent.^4,6,14,17-19^ Furthermore, CSI with tumor boost, a critical prognostic modality for patients with embryonal tumors including PB, is not an option for infants due to adverse neuro-cognitive effects in survivors.^20^ The benefit of high-dose chemotherapy with autologous stem cell rescue versus standard-dose chemotherapy is also unknown. Importantly, the impact of specific treatment on patient survival in the context of the newly recognized molecular subgroups of PB has yet to be evaluated.^21,22^

To address these important and outstanding clinical questions, we assembled and examined a large multi-national cohort of patients with molecularly characterized pineal parenchymal tumors. DNA-methylation profiling was used to exclude alternative CNS tumor diagnoses with histologic resemblance to pineal parenchymal tumors and to distinguish PPTIDs from PBs. Complementary bioinformatic approaches were leveraged to define consensus molecular subgroups and reconcile previously proposed subentities.^5-7^ The landscape of genomic alterations, clinical and treatment factors, patient outcome, and pattern of failure were comprehensively assessed.

## Methods

### Study design and patient treatment

This study encompassed an international meta-analysis of children and adults with pineal parenchymal tumors from three collaborative networks led by the German Cancer Research Center (DKFZ; n=134), the Rare Brain Tumor Consortium/Hospital for Sick Children (RBTC/HSC; n=69), and St Jude Children’s Research Hospital (SJCRH; n=41). With the exception of six new patients from the SJCRH cohort, all others have been reported in separate prior studies.^5-7^ Demographic features, clinical and histologic findings, treatment and patient outcome were annotated and updated based on data availability. Extent of resection was classified as gross-total resection (GTR), near-total resection (NTR), subtotal resection (STR), and biopsy only (Bx). Treatment regimens were summarized according to use, intensity and extent of radiotherapy and/or chemotherapy. Chemotherapeutic protocols involving autologous stem-cell rescue were considered as being high-dose chemotherapy regimens, and all others as standard-dose regimens. Clinical data and tumor material were obtained according to local ethical and Institutional Review Board approval.

### Genome-wide DNA methylation profiling and analysis

FFPE or frozen tumor-derived DNA was profiled on the Infinium HumanMethylation450 (450K) or EPIC BeadChips (Illumina) after bisulfite conversion (Zymo EZ DNA Methylation kit) and as previously described.^5-7^ DNA methylation data and statistical analysis were performed in R version 3.6.3 (www.R-project.org). Raw data were preprocessed using the *minfi* package (Bioconductor), filtered to remove probes on sex chromosomes or those which contained single nucleotide polymorphisms (SNPs), or were not uniquely mapped allowing for one mismatch.^23^ Probes shared between the 450K and EPIC arrays were used for all further analyses. DNA methylation profiles from all samples were compared to a large CNS tumor reference cohort (n>40,000, DKFZ) using t-stochastic neighbor embedding (t-SNE) visualization to exclude molecular outliers, and methylation array-based SNP genotyping was used to identify duplicate samples among the different cohorts.

To define tumor subgroups in the resulting combined cohort, the top 5,000 to 15,000 variably methylated probes based on standard deviation were used in unsupervised cluster analyses by t-SNE, uniform manifold approximation and projection (UMAP), and nonnegative matrix factorization (NMF).^24-26^ Further analyses by k-means and hierarchical clustering (HC, *ConsensusClusterPlus* package v1.44.0) were applied to resolve ambiguous sample membership assignments.^27^ Samples that remained unstable in subgroup membership across multiple methods (n=3) were excluded from downstream analyses. For t-SNE, default parameters were used except for perplexity=10, max_iter=10,000 *(Rtsne* v0.15). For UMAP, default parameters were used except n_epochs = 5,000 *(umap* v0.2.2). NMF analysis was performed with ranks (k) 2-10 at 100 runs *(NMF* v0.20.6). For HC, 1-Pearson correlation for distance measuring and average linkage was used; other settings were default except maxK=10 and reps=10001. k-means was performed with Euclidean for distance measuring and average linkage, with other settings default except maxK=10 and reps=10001. To determine focal and broad chromosomal copy number variations (CNVs), the *conumee* package was used to generate individual copy-number profiles that were validated manually, and composite copy-number profiles per subgroup were assembled based on pre-set thresholds (cut-off for gains=0.15, losses=0.2).^28^

### Genomic and transcriptomic profiling

Somatic and germline driver mutations were studied through whole exome sequencing (WES) or panel-based sequencing as previously described.^5-7^ Specifically, tumor-only sequencing was performed on samples from the DKFZ cohort using an updated 160-gene version of a targeted neuro-oncology panel^29^; tumor-only or paired germline-tumor sequencing for the RBTC/HSC cohort was performed using a combination of whole-exome sequencing (WES) (Genome Quebec or The Centre for Applied Genomics) and targeted sequencing with a six-gene panel for *RB1* and miRNA-processing genes (*DICER1, DROSHA, DGCR8, XPO5, TARBP2*) or a more restricted panel for *DICER1* and *TP53* (ResourcePath, Sterling, VA); while tumor-only or paired WES was performed for the SJCRH cohort. Filtered variants were compiled and frame-shifting or truncating variants (FTVs) in *DICER1, DROSHA, DGCR8* and *RB1*, as well as hotspot missense mutations in the RNase IIIb domain of *DICER1*, were further curated to ensure stringency. In samples molecularly classified as PPTID, sequencing read alignments were manually inspected for exon 4 insertions in *KBTBD4*. Tumor allele frequencies were interpreted in conjunction with methylation-based CNV analysis to determine loss-of-heterozygosity (LOH).

Normalized gene matrices from all samples were combined for transcriptomic analysis. HC was performed based on the top 100 most variably expressed genes to determine transcriptomic similarities among samples. Expression boxplots were created by taking the log2(Normalized counts+1) and likelihood ratio test for differences in expression among subgroups. Differential expression and gene-set enrichment analysis (GSEA) was performed as previously described.^6^

### Clinical correlative and statistical analysis

Continuous variables were expressed by medians and ranges, while categorical variables were summarized by frequencies and percentages. Comparison of variables among subgroups was performed by Fisher’s exact and Wilcoxon rank-sum tests. For survival analyses, patients with incomplete treatment information on use of adjuvant therapy were excluded in analyses related to chemotherapy and radiotherapy use. The date of diagnosis was defined as the date of first biopsy or resection. Overall survival (OS) was defined as the duration between the date of diagnosis and date of either death from any cause or last follow-up; progression-free survival (PFS) was defined as the duration between the date of diagnosis and date of either progression, relapse, death from any cause, or last follow-up. Survival comparisons were performed via log-rank testing.

## Results

### Consensus molecular subgrouping in pineal parenchymal tumors

After exclusion of molecular outliers and genotype duplicates, methylation profiles from 224 patients were used to determine consensus molecular subgrouping (Figure 1A-B). Implementation of dimensionality reduction and clustering analyses resulted in robust assignment of 221 patient samples to five main subgroups (99%, Figure 1C, Supplementary Figure 1 and Supplementary Table 1); three patients with unstable subgroup membership were excluded from subsequent genomic and clinical analyses. We designated the five consensus subgroups, which largely aligned with independent studies (Figure 1D), as PB-miRNA1, PB-miRNA2, PB-MYC/FOXR2, PB-RB1, and PPTID, respectively comprising 43% (96), 10% (23), 15% (34), 11% (25), and 19% (43) of the 221 samples. In keeping with previous report, we observed further methylation subclusters within the PB-miRNA1 (referred to as PB-miRNA1a and PB-miRNA1b) and PPTID (PPTID-a and PPTID-b) subgroups, however no specific genomic or clinical features were associated with these subclusters (Figure 1C and Supplementary Table 2).^7^ Histopathologic grading of tumors designated as CNS embryonal tumors (PB/primitive neuroectodermal tumor [PNET)/trilateral retinoblastoma, Grade 4) or PPTID (Grade 2-3) correlated with molecular classification as PB or PPTID for the majority of samples (Table 1 and Supplementary Figure 2), but 14% (3/21) and 11% (20/189, 16 PB, 4 PNET) of tumors with histology reported as PPTIDs or CNS embryonal tumors had discrepant molecular classification. Two samples with histologic labels of pineal anlage tumors segregated within the PB-MYC/FOXR2 subgroup.

**Table 1.**
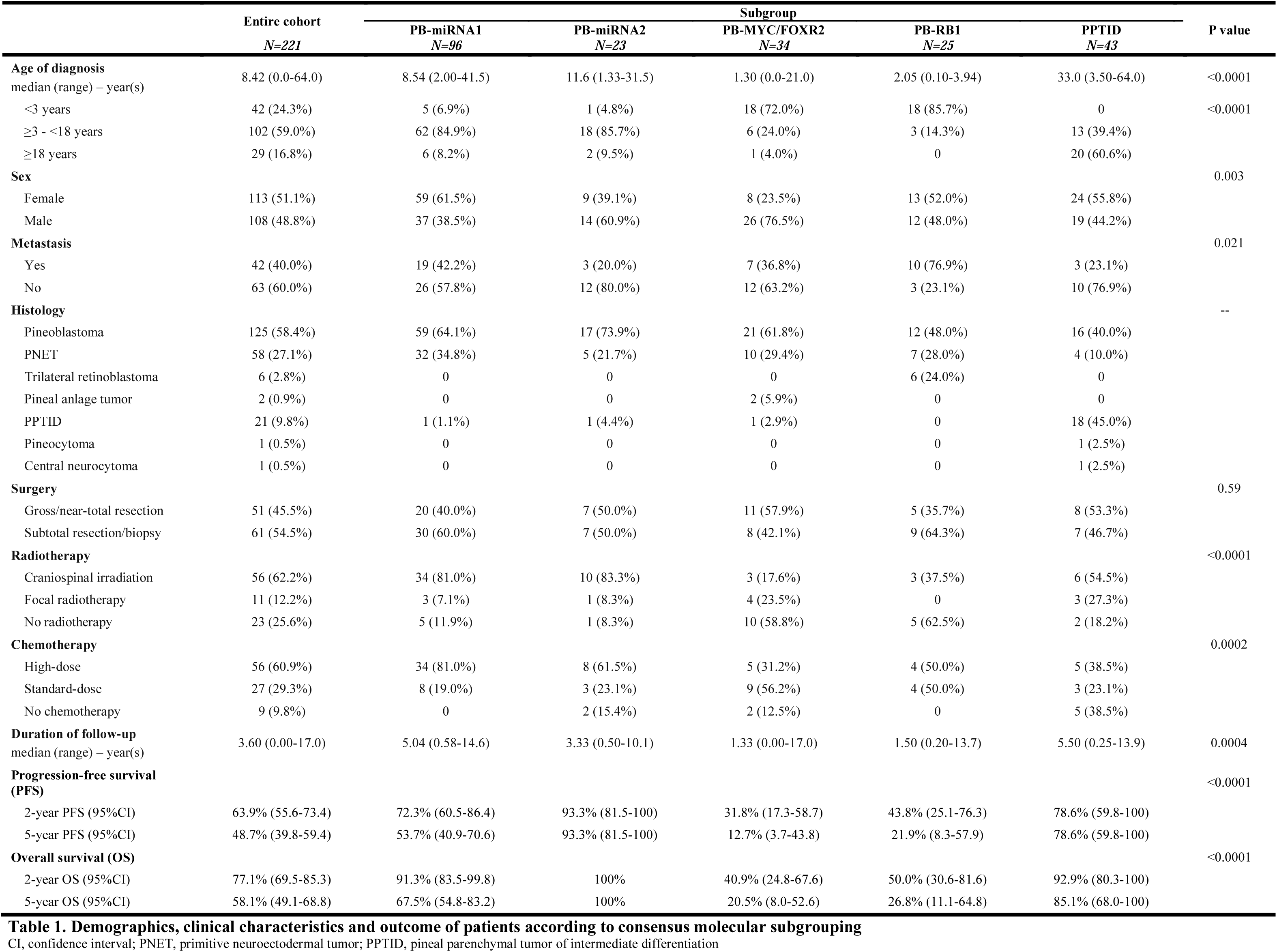
Demographics, clinical characteristics and outcome of patients according to consensus molecular subgrouping. CI, confidence interval; PNET, primitive neuroectodermal tumor; PPTID, pineal parenchymal tumor of intermediate differentiation

**Figure 1.**
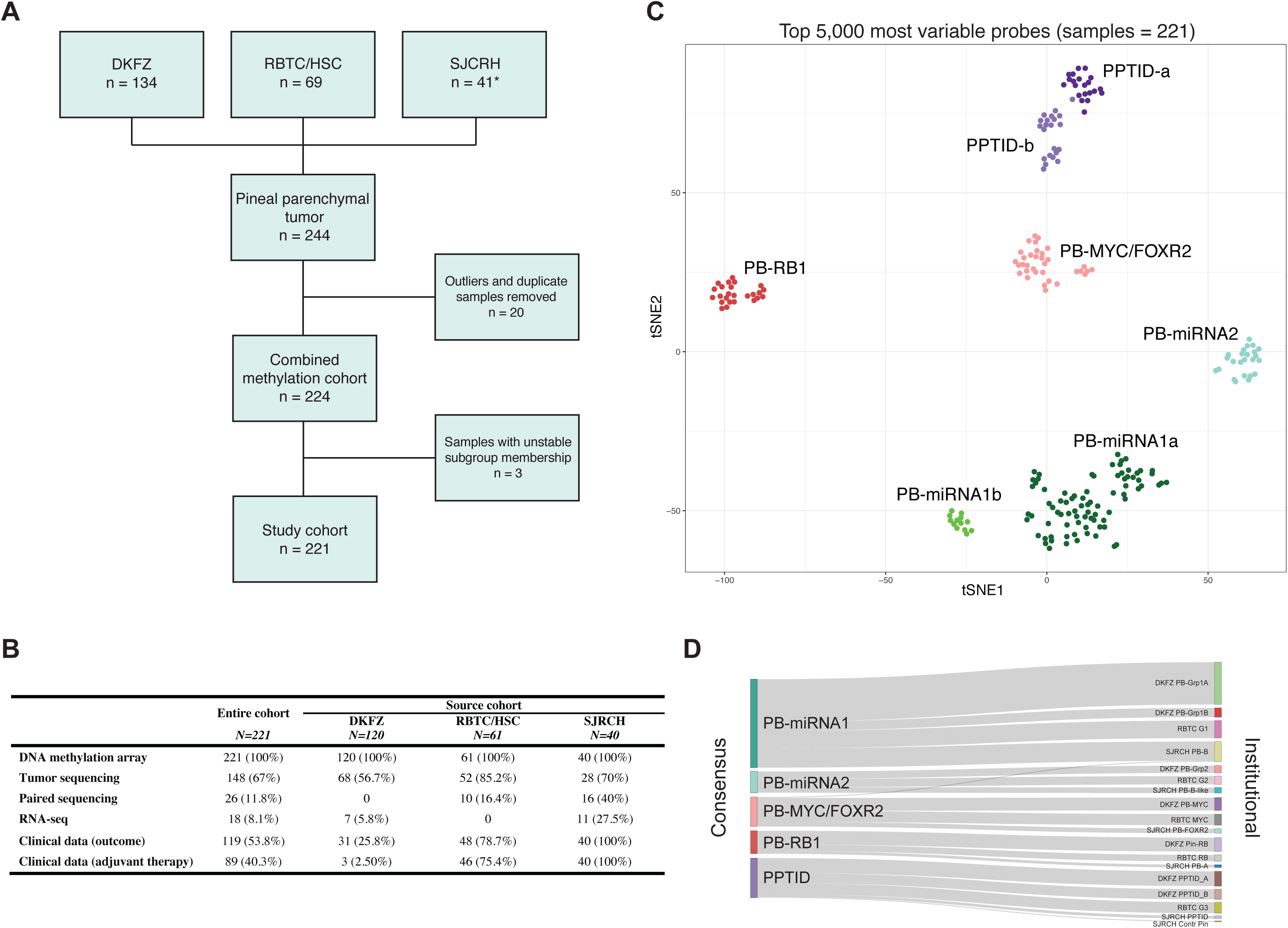
(A) Composition of the consensus study cohort (* including six new patients), (B) availability of genomic and clinical data. (C) UMAP representation of consensus molecular subgroups of pineal parenchymal tumors. (D) Relationship between consensus methylation classes and original institutional designations. DKFZ, German Cancer Research Center; RBTC/HSC, Rare Brain Tumor Consortium/Hospital for Sick Children; SJCRH, St. Jude Children’s Research Hospital.

Although pineal tumors, particularly PB, has generally been considered a disease of younger children, 59% of our cohort was between ≥3-18 years of age, children <3 years represented 24% of cases, while adults (≥ 18 years) represented 17% of patients in our combined cohort. However, patient demographics and disease features were significantly different across the five consensus subgroups (Table 1). PB-miRNA1 or PB-miRNA2 patients were more commonly older children or young adolescents (median age at diagnosis: 8.5 years and 11.6 years, respectively), while patients with PB-MYC/FOXR2 and PB-RB1 were younger children (median age at diagnosis: 1.3 years and 2.1 years, respectively) (Fisher’s exact p<0.0001). Sixty-one percent of molecularly defined PPTIDs were adults with a median age of 33 years.

With the absence of sex predilection (female:male=113:108; 51%:49%) across the entire cohort, 61% of patients in the PB-miRNA2 subgroup and 77% of patients in the PB-MYC/FOXR2 subgroup were male (Fisher’s exact p=0.003). Metastatic status also differed significantly between tumor subgroups (Fisher’s exact p=0.021). Patients from the PB-miRNA1, PB-miRNA2, PB-MYC/FOXR2 and PPTID subgroups more likely presented with localized disease (42%, 20%, 37% and 23% demonstrating metastasis at diagnosis respectively). In contrast, 77% of patients from the PB-RB1 subgroups had metastatic disease at diagnosis.

### The genomic landscape of pineal parenchymal tumors

Tumor molecular subgroups were enriched for distinct driver gene alterations and broad chromosomal copy-number variations (CNVs) (Figure 2). Overall, 65% (53/81) of sequenced tumors from the miRNA subgroups had mutually exclusive alterations of miRNA-processing genes. Among 60 PB-miRNA1 samples sequenced, FTVs and/or focal losses of *DICER1, DROSHA*, and *DGCR8* were observed in 16 (27%), 15 (25%), and 5 (8%) cases, respectively; additional samples with focal *DROSHA* (n=7, 19%) and *DGCR8* (n=2, 6%) loss were identified by copy-number analysis in cases without sequencing data (n=36). PB-miRNA1 tumors also had frequent arm-level gains of chromosome 7, 12, and 17. Amongst 21 PB-miRNA2 subgroup samples with sequencing data, we observed *DICER1* and *DROSHA* alterations respectively in 11 (52%) and 6 (29%) tumors while 74% had chr 14q loss. Comparison between *DICER1-*altered PB-miRNA1 and 2 tumors did not reveal differences in the genetic region affected by mutations (Figure 2C). However, while *DICER1* FTVs in PB-miRNA2 tumors were always accompanied by chr 14q loss (presumably leading to complete loss-of-function), PB-miRNA1 tumors most commonly exhibited two different FTVs, or FTV with apparent copy-number neutral LOH (Figure 2D). Notably, paired-sequencing data available showed that 6/12 patients with *DICER1*-altered PBs had *DICER1* mutations that were germline in origin, and were encountered in both PB-miRNA1 and miRNA2 subgroups. In contrast to other *DICER1* -associated CNS and extra-CNS tumors, no hotspot missense mutations in the *DICER1* RNase IIIb domain were present in our cohort.^10,30^

**Figure 2.**
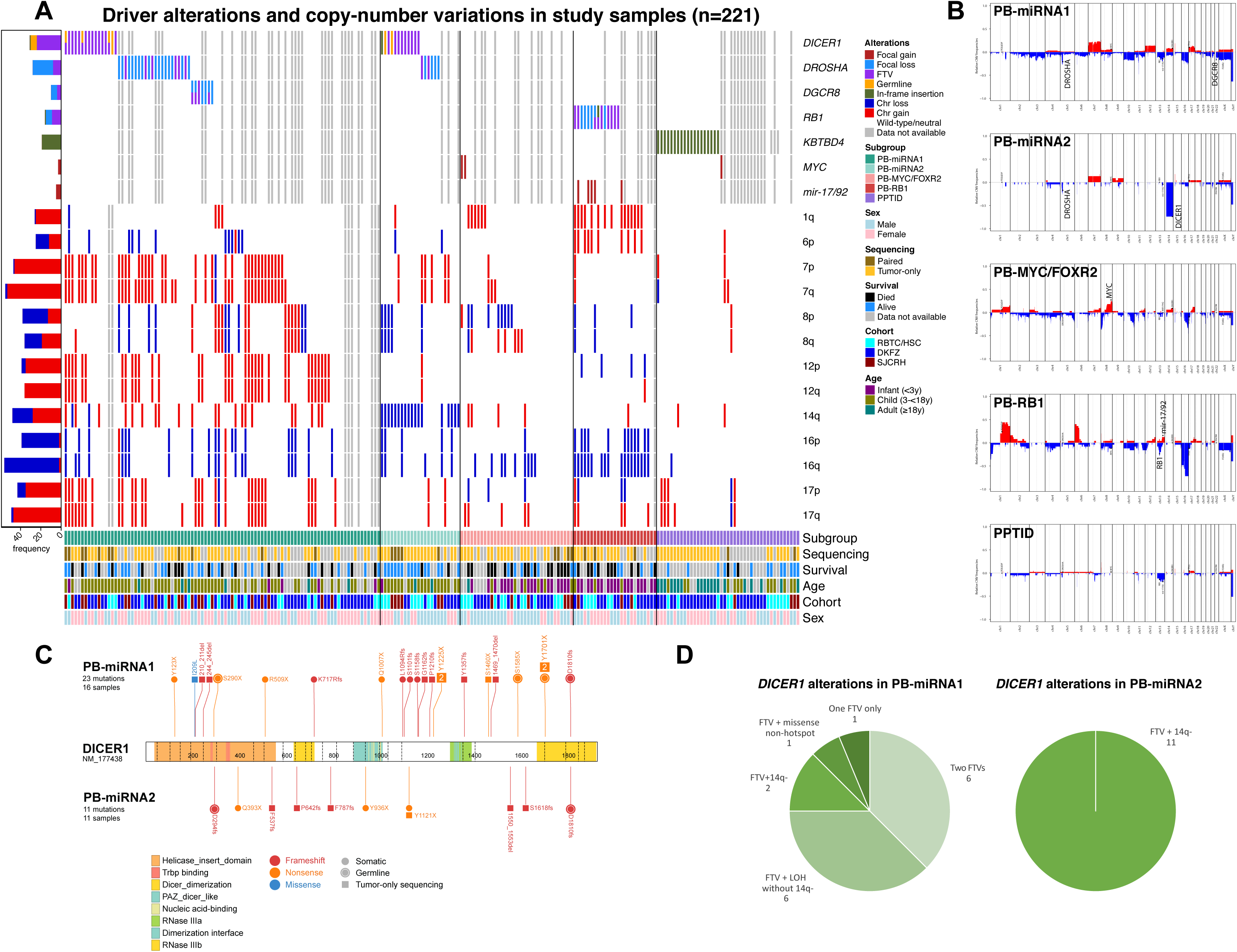
Recurrent genomic events in pineal parenchymal tumors. (A) Oncoprint depicting nature of driver alterations and recurrent copy-number variations (CNVs) by methylation subgroup. (B) Composite genome-wide CNV plots by methylation subgroups. (C) Comparison of *DICER1* variants and (D) types of *DICER1* alterations identified in PB-miRNA1 vs. PB-miRNA2 tumor samples. chr, chromosome; DKFZ, German Cancer Research Center; FTV, frameshifting or truncating variant; RBTC/HSC, Rare Brain Tumor Consortium/Hospital for Sick Children; LOH, loss of heterozygosity; SJCRH, St. Jude Children’s Research Hospital; y, year(s)

No recurrent genetic driver mutations were seen in 23 PB-MYC/FOXR2 tumors sequenced (Figure 2). CNV analyses showed common broad chr 8p and 16q loss and focal 8q amplification targeting *MYC* in two samples (6%), while RNA-seq data available for four tumors indicated subgroup-specific over-expression of the *FOXR2* proto-oncogene (Figure 3 and Supplementary Figure 3). Seventy-five percent of PB-RB1 tumors sequenced (12/16) harbored FTV (n=7), focal deletion (n=4), or a combination of deleterious events (n=1) involving *RB1*. Focal gain of *miR-17/92* was observed in four (25%) samples sequenced including three with *RB1* alteration. Of nine PB-RB1 samples without sequencing data, one each respectively carried focal *RB1* loss and *miR-17/92* gain. Similar to observations in retinoblastoma samples, chr 1q and 6p gain and chr 16 loss were common cytogenetic events in PB-RB1.^31^ Six patients (24%) from this subgroup had a clinical diagnosis of trilateral retinoblastoma considered pathognomonic for *RB1* predisposition syndrome (Table 1 and Supplementary Figure 2). In the PPTID subgroup chromosomal CNVs were infrequent, however 73% of sequenced samples (19/26) had in-frame insertions of *KBTBD4*. Adult patients (≥18 years) in the PPTID cluster were more likely to have *KBTBD4-mutant* tumors than patients <18 years of age (14/16 vs. 3/7, Fisher’s exact p=0.045, Supplementary Table 3).

**Figure 3.**
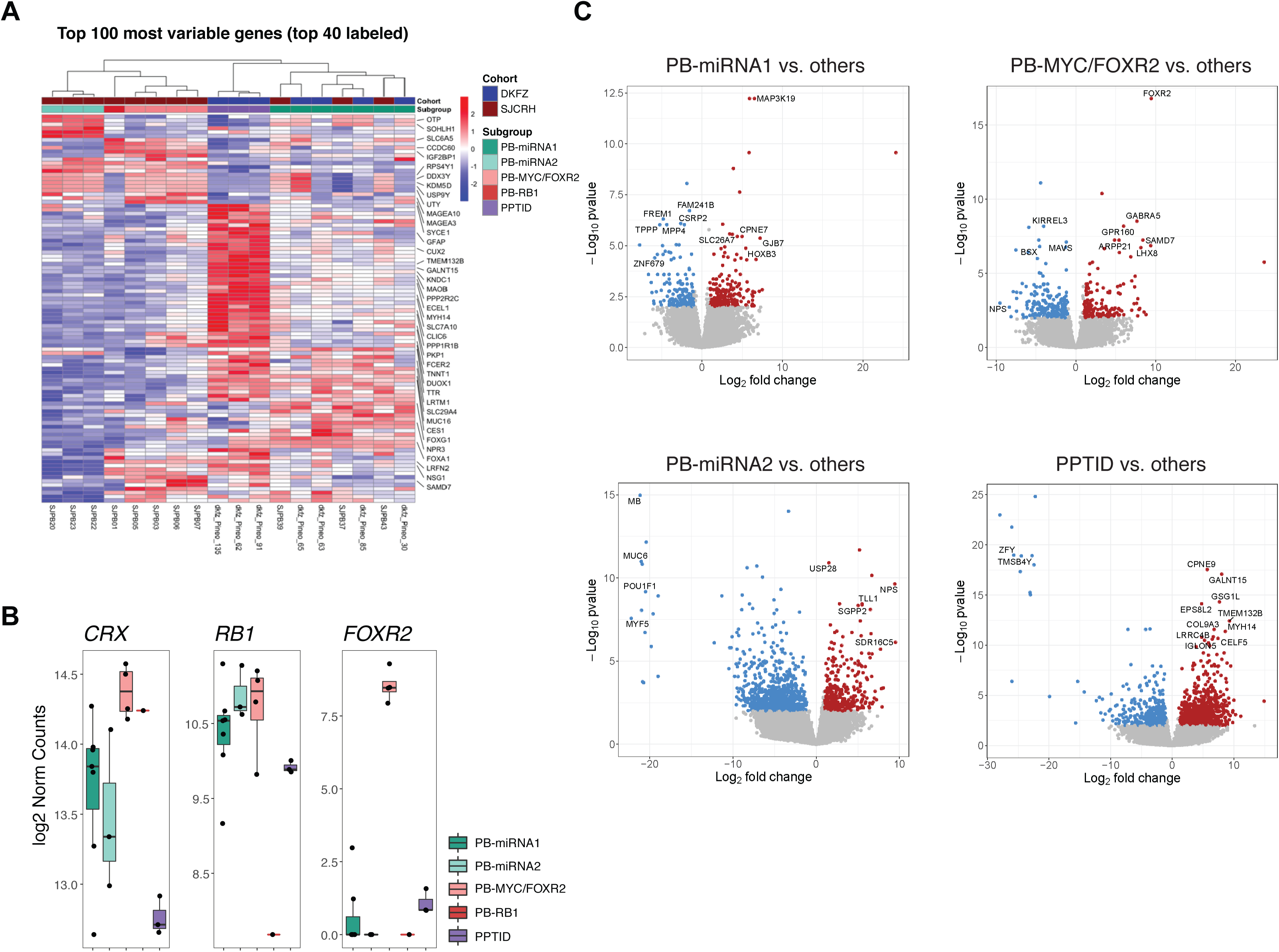
Transcriptomic analysis of study samples (n=18). (A) Clustering based on top 100 differentially expressed genes. (B) Comparison of expression level of *CRX* - canonical pinealocyte marker, *FOXR2, RB1* among tumor subgroups. (C) Differential expression analysis between tumor from specific subgroups and other samples. DKFZ, German Cancer Research Center; SJCRH, St. Jude Children’s Research Hospital

Clustering analyses of available gene expression data (n=18) recapitulated the molecular subgrouping derived from methylation analysis (Figure 3). High *CRX* expression was observed across subgroups supporting their pineal parenchymal origins. GSEA based on limited RNA-seq data available revealed enrichment of “immune-related” and “positive regulation of ERK1 and ERK2 cascade” gene-ontology (GO) terms with significant overexpression of *MAP3K19* in PB-miRNA1 tumors, while enrichment of the “phototransduction” GO term was observed in PB-MYC/FOXR2 tumors.

### Clinical and treatment related prognostic features for patients with pineal parenchymal tumors

Reflecting the challenge of surgery for pineal region tumors, less than half of patients with surgical data underwent GTR/NTR (51/112, 46%). More than half of patients (56/90, 62%) with radiation treatment data received CSI, while 11 (12%) and 23 (26%) respectively had focal or no radiotherapy. Amongst 92 patients with chemotherapy detailed, 56 (61%), 27 (29%) and 9 (10%) of patients respectively received high-dose, standard-dose or no chemotherapy.

Complete data on adjuvant therapy and survival were available for 89 (40%) and 119 (54%) patients respectively (Figure 1B). The median duration of follow-up was 3.6 years (range 0-17) for all patients and 5.5 years (range 0.3-17) for surviving patients. Overall, PFS and OS rates were significantly different across the molecular subgroups (PFS and OS log-rank p<0.0001, Figure 4A-B). Patients with PB-miRNA2 had superior 5-year PFS and OS of 93.3% (95% confidence interval [CI]: 81.5-100) and 100%, respectively, while PB-miRNA1 patients exhibited intermediate outcomes with respective 5-year PFS and OS of 53.7% (95%CI: 40.9-70.6) and 67.5% (95%CI: 54.8-83.2). Overall, patients with PB-MYC/FOXR2 and PB-RB1 had the poorest outcomes with respective 5-year PFS of 12.7% (95%CI: 3.7-43.8) and 21.9% (95%CI: 8.3-57.9) and respective 5-year OS of 20.5% (95%CI: 8.0-52.6) and 26.8% (95%CI: 11.1-64.8). For all patients with molecularly defined PBs, age (PFS and OS log-rank p<0.0001), metastatic status (PFS log-rank p=0.0041; OS log-rank p=0.0031), use and volume of radiotherapy (PFS and OS log-rank p<0.0001), and receipt and intensity of chemotherapy (PFS and OS log-rank p<0.0001) were significantly associated with patient outcome (Figure 4C-F,I-L). For the entire cohort, GTR/NTR, even when stratified by metastatic status, did not offer significant survival advantage (Figure 4G-H and Supplementary Figure 4).

**Figure 4.**
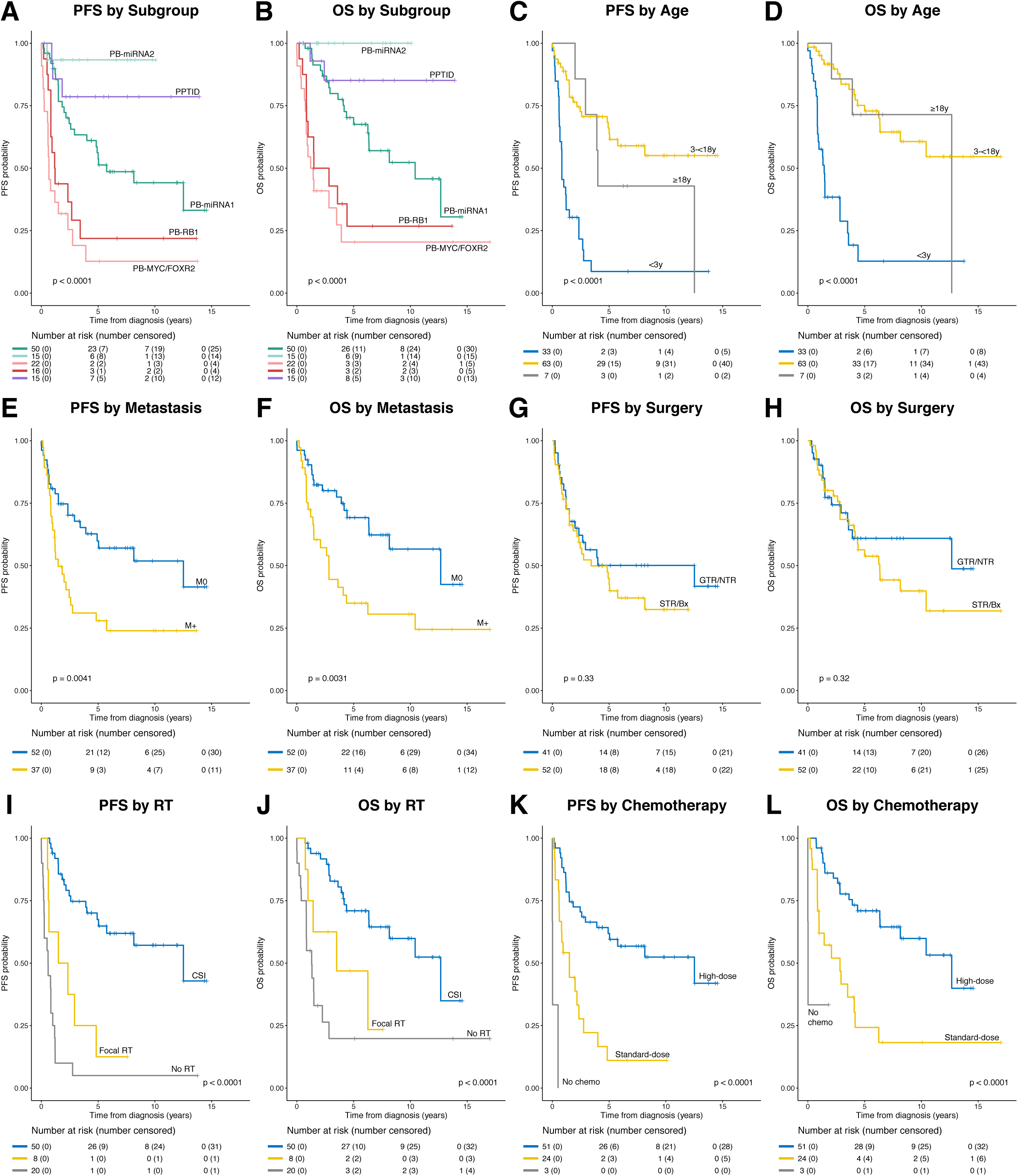
Progression-free (PFS) and overall survival (OS) of (A-B) the entire study cohort by methylation subgroup; and outcome of molecularly defined pineoblastomas according to (C-D) age at diagnosis, (E-F) metastatic status, (G-H) extent of resection, (I-J) radiotherapy (RT) use, and (K-L) chemotherapy (chemo) use. Bx, biopsy; CSI, craniospinal irradiation; GTR, gross-total resection; M0, non-metastatic; M+, metastatic; NTR, near-total resection; STR, subtotal resection; y, year(s)

Analysis by subgroups revealed varied impact of clinical and treatment factors on outcome (Supplementary Figure 5). Metastatic status was significantly associated with PFS and OS in PB-miRNA1 but not in other PB subgroups (PFS log-rank p=0.049; OS log-rank p=0.016). All three patients with metastatic disease and data on outcome in PB-miRNA2 subgroup survived without progression, whereas those with PB-MYC/FOXR2 and PB-RB1 were at risk of progressive disease regardless of disease extent. Among patients with PB-miRNA1, the use and intensity of radiotherapy as well as chemotherapy were significantly associated with PFS and OS. All patients with PB-miRNA2 and data on outcome survived; while the majority received CSI and high-dose chemotherapy, one patient received focal radiotherapy with high-dose chemotherapy, three patients received CSI with standard-dose chemotherapy. The only PB-miRNA2 patient who progressed after receiving surgery alone was salvaged with CSI and standard-dose chemotherapy. PFS in the PB-MYC/FOXR2 and PB-RB1 subgroups was anecdotal. Two survivors in PB-MYC/FOXR2 subgroup with known information on adjuvant therapy received CSI with high-dose chemotherapy, and high-dose chemotherapy alone respectively, while the patient with PB-RB1 received CSI and high-dose chemotherapy. Patients with molecularly defined PPTID had 5-year PFS and OS rates of 78.6% (95%CI: 59.8-100) and 85.1% (95%CI: 68.0-100), respectively. Both patients with metastatic disease died of progression, three patients with localized disease remained in remission after focal therapy with (n=2) or without (n=1) chemotherapy. Among patients in the PB-miRNA1/2 and PPTID subgroups, survival did not differ according to the presence or absence of specific driver alterations (Supplementary Figure 6).

Site of disease progression was annotated in 29 patients, with 21 (72%) suffering from distant failure, five (17%) from combined distant and local failure, and three (10%) with local failure (Supplementary Table 4). The site of failure did not differ significantly by molecular subgroup or radiotherapy use and extent. Seven patients were alive at last follow-up for more than one year after disease progression, and details on second-line therapy were available for five patients, all of whom were salvaged by CSI with chemotherapy in the context of upfront therapy that consisted of focal or no radiotherapy (Supplementary Table 5).

## Discussion

Based on the largest reported molecularly-defined pineal parenchymal tumor cohort, we harmonized previously described inter-tumoral heterogeneity in PB and proposed the nomenclature of consensus subgroups – PB-miRNA1, PB-miRNA2, PB-MYC/FOXR2 and PB-RB1. These molecular subgroups are characterized by critical differences in demographic, clinical and genomic features, and display drastic discrepancies in patient survival. Analysis of the molecularly-defined PB cohort recapitulated known prognostic factors, such as metastatic status, age at diagnosis, and radiation use. Importantly, extent of initial surgical resection was not significantly associated with patient survival, either in the entire molecular PB cohort, stratified by metastatic status, or by subgroup. It is possible that reported benefit of more aggressive resection in previous studies was confounded by higher rates for total tumor removal in patients with localized disease.^6,32^ The recognition of heterogeneity within PB is key to enhancing patient outcome, as the effect of adjuvant treatment could be scrutinized in the context of subgroup tumor biology. Accordingly, treatment intensity could be adapted and use of experimental therapy prioritized.

PBs with altered miRNA-processing pathway, PB-miRNA1 and PB-miRNA2, were more commonly seen in older children and represented eight out of nine cases known to be in the adult age group from our cohort. Despite a reasonably similar treatment approach that incorporated CSI in >80% of patients in either subgroups, stark differences in outcome were observed. All patients in the PB-miRNA2 subgroup with reported outcome survived, including two with metastatic disease, one with localized disease who received focal radiation and chemotherapy. The omission of adjuvant therapy in one patient, however, resulted in early relapse but could be salvaged with a CSI-containing regimen. It is therefore prudent for future studies to prospectively evaluate treatment with reduced-dose CSI in patients with PB-miRNA2. On the contrary, patients with PB-miRNA1 carried an intermediate outcome may benefit from intensive adjuvant therapy comprising of CSI and high-dose chemotherapy. Notably, patients with PB-miRNA1 uniquely exhibited late disease progression, calling for the consideration of maintenance therapy and the need to continue disease surveillance. Mechanisms that underlie the varied disease aggressiveness between these two subgroups remain elusive, although subgroup-specific patterns of miRNA-processing gene alterations were observed. Specifically, PB-miRNA2 had higher rates of tumors with *DICER1* alterations than in PB-miRNA1; and while it is now known that PBs tolerate biallelic inactivation of *DICER1*, we demonstrate that *DICER1* -mutant PB-miRNA2 tumors universally carry whole chr 14q loss as the second-hit mechanism.^30,33^ In two patients sharing the same germline *DICER1* FTV, second FTV versus 14q loss as the somatic hit resulted respectively in tumors with PB-miRNA1 (SJPB26, M3 disease with very high Ki67) and PB-miRNA2 (RBTC803, M0 disease with 10-15% Ki67) signatures. It might be speculated that the nature of the secondary events leading to complete loss of *DICER1* function could dictate disease course in PBs. In view of the upregulation of immune-related GO terms in PB-miRNA1, subgroup-specific tumor microenvironment and possible immune infiltrates might be exploited as therapeutic vulnerabilities in future clinical trials.

Forty-percent of children less than three years with molecularly-defined PB belonged to the PB-MYC/FOXR2 subgroup and another 40%, the PB-RB1 subgroup. Outcome for patients from these subgroups was dismal, with rapid progression observed. Such inferior survival might have been associated with the inability to administer CSI and general reluctance in radiotherapy use in these very young children.^4^ Notwithstanding the theoretical benefit of CSI, the short time-to-progression implies that radiotherapy deferral approaches might be impractical. Furthermore, extended irradiation fields pose an increased risk for subsequent malignancies in patients with germline *RB1* mutations.^34^ *In vitro* analyses have demonstrated the cooperative role between *FOXR2* and *MYC* through direct interaction and complex formation, in turn promoting *MYC* transcriptional activities, cellular proliferation and tumorigenesis.^35^ Although further studies to confirm *MYC* dependency in PB-MYC/FOXR2 tumor without *MYC* amplification are warranted, experimental approaches that might be explored include BET-bromodomain inhibition which have shown preclinical activity against MYC-driven medulloblastomas.^36,37^ For patients with PB-RB1, *in silico* drug screening based on a recently established R6-deficient murine PB model suggested potential activity with nortriptyline, an anti-depressant, through lysosome disruption.^38^ Further research on treatment strategies for these disease subgroups is urgently needed. With two-third of PB-FOXR2/MYC patients presenting with localized disease, the efficacy of upfront focal radiotherapy, especially with proton beam, followed by high-dose chemotherapy could be evaluated while awaiting novel therapeutic agents.

PPTIDs are molecularly and clinically distinct from PBs, although the histologic delineation was not straightforward in up to 14% of cases. Previously proposed “Group 3” PBs that harbored *KBTBD4* insertions are aligned with PPTIDs in our meta-analysis cohort.^5^ In clinical practice, targeted evaluation for disease-defining driver genes might facilitate diagnosis in cases with indeterminate histomorphology.^39^ Such ambiguity also represents a caveat in the interpretation of clinical studies on pineal parenchymal tumors especially in adolescents and adults, where both entities are encountered. In our study, patients with molecularly-defined PPTIDs had satisfactory outcome with 13 out of 15 patients surviving without disease. Both patients who died of disease had metastasis at presentation. Among patients with details on treatment regimen, three patients who received focal radiotherapy were survivors at 3.2-5.5 years from diagnosis. Previous studies have demonstrated the prognostic value of adjuvant radiotherapy use in histologically diagnosed PPTIDs, with little being known regarding the impact from field of treatment; while the benefit of adjuvant chemotherapy has not been established.^39,40^ We suggest that a prospective trial for patients with localized, molecularly-confirmed PPTIDs (by methylation profiling and/or detection of *KBTBD4* insertion) to be treated with focal radiotherapy with or without standard-dose chemotherapy should be undertaken.

Our study is inherently limited by its retrospective design and non-uniformity in treatment strategy adopted. The lack of central pathologic review did not permit systematic correlation of histologic findings, such as proliferative labeling index, with molecular results, nor did it allow validation of genomic findings by orthogonal tools. Due to the rarity of pineal parenchymal tumors, however, the unprecedented clinical-molecular cohort is of value for allowing precise description of clinical and genomic features, as well as appreciation of the influence of treatment factors within harmonized disease subgroups. In conclusion, we corroborated the clinical and biological heterogeneity within pineal parenchymal tumors and proposed consensus molecular subgroups. Future PB clinical studies should stratify patients molecularly to allow personalized management.

## Data Availability

Genomic data will be available from the authors upon reasonable request.

## Acknowledgement

The authors would like to acknowledge the clinical and research staff of participating sites for contributing to the care and study of all involved patients.

